# Implementing the SWYC: A Quality Improvement Project and Its Impact on Care Delivery

**DOI:** 10.1101/2025.01.28.25320926

**Authors:** Yusuke Matsuura, Felix Richter, Gabrielle Block, Carolyn Rosen, Cynthia Katz

## Abstract

**Background:** Despite AAP recommending the use of validated developmental screening tools, national adoption remains low. To address this issue, we initiated a QI project to implement the Survey of Well-being in Young Children (SWYC) and assess its impact within our residency clinic. Our objective was to achieve over 50% screening coverage for well-child visits for children aged 8 to 33 months, with equitable implementation for both English-primary and non-English-primary speaking families. Additionally, we hypothesized that SWYC would significantly increase early intervention (EI) referral rates.

**Methods:** Patients aged 8 to 33 months attending well-child visits were included. During implementation, various interventions, including staff emails, daily huddles, and integration of SWYC into the electronic health record, were employed. As part of the clinic workflow, a designated social worker (SW) initiates EI referrals; thus, SW referrals served as a proxy for EI referrals. Referral rates were compared pre- and post-SWYC implementation. Statistical significance was assessed using a chi-square test in R software.

**Results:** SWYC utilization reached 50% within five months of implementation. Analysis showed no significant difference in SWYC use between English-primary and non-English-primary speakers (p = 0.13). Post-SWYC, SW referral rates increased by 34% compared to pre-SWYC rates (p = 0.009).

**Conclusions:** This QI project showcased the swift adoption of a standardized developmental screening tool through system and cultural changes. SWYC implementation significantly increased SW referral rates, suggesting improved identification of developmental delays. Standardized developmental screening has the potential to enhance developmental outcomes and promote care equity through objective assessment systems.

## Introduction

The prevalence of developmental disabilities has been increasing overtime, with approximately one in five children aged 3 to 17 having a disability, such as attention-deficit/hyperactivity disorder (ADHD), autism spectrum disorder (ASD), or an intellectual disability.^1^ Developmental disabilities also include delays in physical, cognitive, social/emotional, communication, and adaptive domains, as well as other disorders associated with congenital and genetic conditions.^2^ The exact reason for this rise remains unclear; however, early detection and intervention of developmental disabilities are becoming increasingly important for children’s health and future outcomes.^3-7^ The responsibility for early detection largely falls on general pediatricians, who are expected to integrate screenings into their already busy well-child visits. Without the use of a standardized tool, detection relies solely on the clinical acumen of the provider, which can be influenced by many factors such as personal provider experience, parental recall, time constraints, and external practice flow issues. The AAP has recommended performing developmental screening using a validated screening tool during the 9-, 18-, and 30-month well-child visits,^8,9^ and AAP guidelines have led to an increase in overall screening, with rates increasing from 23% in 2002 to 63% in 2016.^10,11^ However, rates remain subpar nationally.^12^ Mount Sinai Kravis Children’s Hospital, Cohen Center for Pediatric Comprehensive Care is an urban General Pediatrics academic practice located in the Upper East Side/East Harlem neighborhood of Manhattan in New York City.^13^ The site functions as the main continuity clinic site for the Pediatric Residency Program at the Icahn School of Medicine at Mount Sinai.^14^ It is critical for residents to receive training in the detection and management of developmental delays during clinic encounters before launching into independent practice.^15^ Prior to this project, the developmental screening for our patients consisted of age-appropriate surveillance questions taken from a variety of screening tools but not validated as a stand-alone assessment tool in our Electronic Health Record (EHR).

We set out to implement a process to standardize our assessment of developmental milestones by using a validated screening tool to meet the standard of care recommended by the AAP, as well to optimize the education around developmental assessment and referrals. We anticipated that the implementation of a standardized screening process would improve our ability to identify potential developmental delays in a timely manner, refer to appropriate services, and improve the long-term outcomes for our patients.

## Objectives

We implemented a Quality Improvement (QI) initiative in April 2022 to address a gap in care in our ambulatory general pediatrics clinic. The primary goal was to screen at least 50% of patients attending well-child visits between 8 months and 33 months (corrected for prematurity up to 24 months) using a validated developmental screening tool by June 2024. This age range was chosen to align with the eligibility criteria for Early Intervention (EI) services. Additionally, the initiative aimed to ensure equitable screening practices for both English-speaking patients and non-primary English-speaking patients. As a secondary outcome, we aimed to determine whether the implementation of a standardized screening tool leads to an increase in referral rates to EI services.

## Methods

### Setting

This QI project was conducted at the Cohen Center for Pediatric Comprehensive Care, the pediatric resident continuity clinic site of The Mount Sinai Hospital in New York City. We are a medium sized pediatric residency program consisting of primarily categorical pediatric residents as well as residents in combined training programs including child neurology, pediatric genetics, and triple board (pediatrics, psychiatry, and child psychiatry). The catchment area for patients who are followed at the Cohen Center for Pediatric Comprehensive Care includes all 5 boroughs of NYC: Bronx, Manhattan, Queens, Brooklyn, Staten Island, and broader areas including Westchester and New Jersey. The practice conducts approximately 2,000 well-child and follow-up visit monthly, or which approximately 200 per month included our study’s target age groups. Our study excluded urgent care visits.

### Project Timeline Overview

Our developmental screening QI project team consisted of thirteen residents and two supervising attending. During the project’s first four months (July 2022 – October 2022), the QI team focused on a literature search for selecting and implementing a validated developmental screening tool. We also defined our global aim and utilized multiple QI tools during the project planning process. The process map visualized each step and clarified how implementation should occur within the practice workflow.^16,17^ A fishbone diagram was used to identify potential barriers to implementing the SWYC screening in the clinic.^18,19^ A key driver diagram helped identify appropriate and feasible interventions to achieve our aim.^20,21^ From November 2022 through early February 2023, we implemented the Survey of Well-being of Young Children (SWYC) in EHR and initiated data collection. From mid-February 2023 onward, we disseminated information about SWYC implementation to all clinicians followed by iterative Plan-Do-Check-Act (PDCA) cycles of continuous data collection and analysis. This project was approved and deemed to be QI the Mount Sinai Kravis Children’s Hospital QI Performance Committee. As per the committee’s decision, IRB approval was determined to be unnecessary.

### Literature Review and Determination of Implemented Tool

A comprehensive literature search was conducted using PubMed, to inform our choice of a validated screening tool for implementation and resulted in the reviewing more than 30 papers. The study titled “Comparative Accuracy of Developmental Screening Questionnaires,” published in JAMA,^22^ was particularly helpful in assessing the specificity and sensitivity of three commonly used developmental screening instruments for children: the Ages and Stages Questionnaires, Third Edition (ASQ-3); the Survey of Well-being of Young Children (SWYC); and the Parents’ Evaluation of Developmental Status (PEDS). The study reported specificities for younger children (0-42 months) of 89.4%, 89.0%, and 79.6%, respectively, and sensitivities for severe delays of 60.0%, 73.7%, and 78.9%. Both the ASQ-3 and SWYC Milestones demonstrated higher specificity than the PEDS (P < 0.001 and P = 0.002, respectively). However, differences in sensitivity were not statistically significant. Sensitivity exceeded 70% only in cases of severe delays, with the SWYC and the PEDS among younger children. After evaluating the content, time required for completion, and feasibility of implementation into our system, the SWYC was chosen for use. Notably, among these three screening tools, only the SWYC is available free of charge.

The SWYC, developed by a team of pediatricians and researchers at Tufts Medical Center, was designed to be a comprehensive, freely available developmental screening tool for use in pediatric primary care settings.^23,24^ The SWYC covers various aspects of a child’s development and well-being, such as motor skills, language, and social-emotional development, as well as parental concerns about behavior, emotions, and family stressors. For this QI initiative, we decided to focus our use of the SWYC only on the developmental milestones’ questionnaire section. The developmental milestones questionnaire section of the SWYC poses 10 questions intended for caregivers to assess a child’s skills across language, gross motor, fine motor and personal-social domains. Caregivers are asked to assess a child’s readiness as “not yet (0),” “somewhat (1),” or “very much (2)” A score is then calculated indicating whether the child “appears to meet age expectations” or “needs review”. (Table 1). Figure 1 is an example of the SWYC questionnaires for 9-month-olds, along with the score interpretation sheet (Figure 2).

**Table 1:** Inc Included Age Milestones in the SWYC Form.

| **SWYC FORM** | **INCLUDED MONTHS** |
| --- | --- |
| 2-MONTH FORM | (1 month, 0 days to 3 months, 31 days) |
| 4-MONTH FORM | (4 months, 0 days to 5 months, 31 days) |
| 6-MONTH FORM | (6 months, 0 days to 8 months, 31 days) |
| 9-MONTH FORM | (9 months, 0 days to 11 months, 31 days) |
| 12-MONTH FORM | (12 months, 0 days to 14 months, 31 days) |
| 15-MONTH FORM | (15 months, 0 days to 17 months, 31 days) |
| 18-MONTH FORM | (18 months, 0 days to 22 months, 31 days) |
| 24-MONTH FORM | (23 months, 0 days to 28 months, 31 days) |
| 30-MONTH FORM | (29 months, 0 days to 34 months, 31 days) |
| 36-MONTH FORM | (35 months, 0 days to 46 months, 31 days) |
| 48-MONTH FORM | (47 months, 0 days to 58 months, 31 days) |
| 60-MONTH FORM | (59 months, 0 days to 65 months, 31 days) |

**Figure 1:**
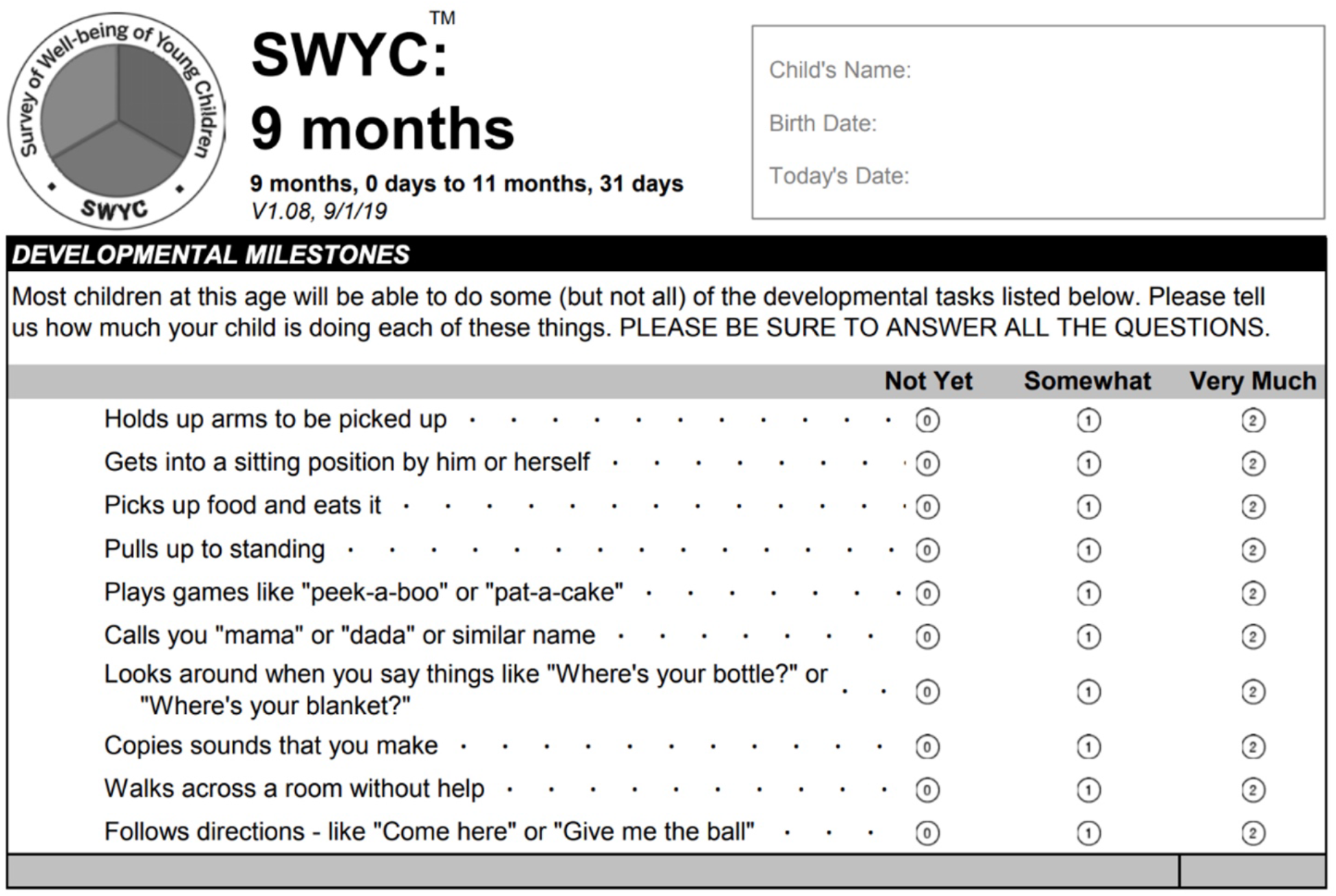
SWYC Questionnaires for 9-month-olds, Developmental Milestones Reproduced from Tufts Medicine, ‘9-Month English Age-Specific,’ available at [https://www.tuftsmedicine.org/sites/default/files/2024-01/9-Month-English-Age-specific-2021.pdf]

**Figure 2:**
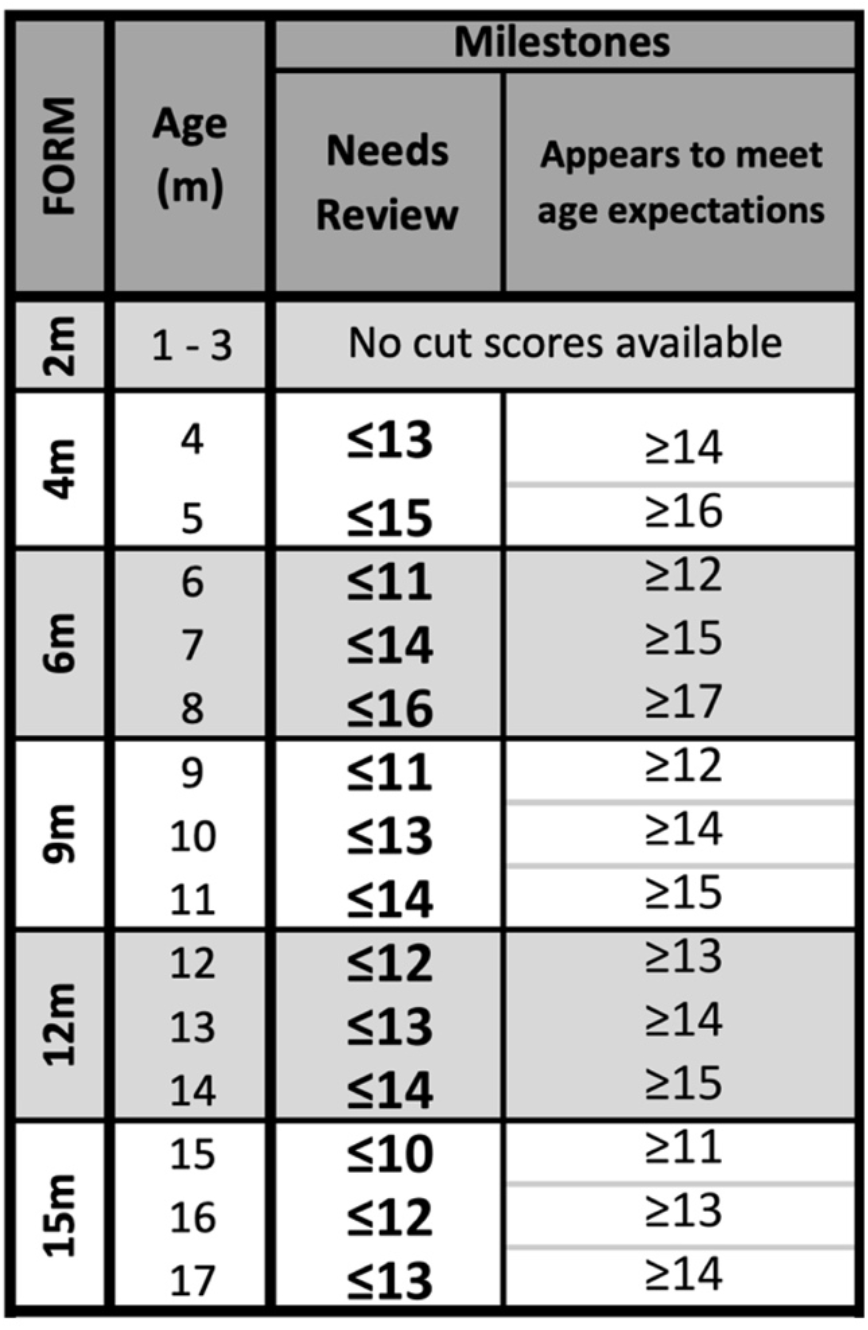
SWYC Score Interpretation Sheet Reproduced from Tufts Medicine, ‘Scoring Cheat Sheet v2,’ available at [https://www.tuftsmedicine.org/sites/default/files/2024-01/scoring-cheat-sheet-v2.pdf].

### Inclusion/Exclusion Criteria

The study population included patients who attended well child visits at the Cohen Center for Pediatric Comprehensive Care between the ages of 8 months – through 33 months (corrected for gestational age through 24 months). Exclusion criteria included patients being seen for urgent visits, well-child visits for patients younger than 7 months or older than 34 months of age.

### Measures

The outcome measure was the percentage of patients aged 8 to 33 months presenting for well-child visits with a completed SWYC questionnaire documented in the EHR. Data were collected via Excel reports generated by the IT team, and rates were compared between primary English-speaking and non-primary English-speaking patients to assess disparities based on preferred language.

The process measure was the percentage of patients who completed a paper SWYC questionnaire before the physician entered the room. Since the Epic EHR platform at the Mount Sinai Health System had not yet implemented an electronic SWYC questionnaire via the patient portal, paper forms in English and Spanish were temporarily distributed at check-in. During the pilot phase in December 2022, several challenges arose, including maintaining questionnaire supplies, incomplete forms, and difficulties identifying eligible visits and assigning age-appropriate forms. These challenges led to the discontinuation of the paper-based system, with a shift in focus to physicians completing the SWYC within the EHR during patient encounters.

The balancing measure aimed to evaluate the potential impacts on pediatric residents’ annual In-Training Examinations (ITE) scores due to reduced active learning of developmental milestones. Embedding the tool in Epic might lead to less internalization of these milestones and potentially lower exam scores. However, this measure was not assessed due to the privacy of ITE results.

### EHR Integration

In order to optimize successful implementation, we decided to seek integration of the SWYC into Epic. With help from our local Epic IT team, the SWYC was integrated into our outpatient encounters. Each questionnaire was mapped to the patient’s corrected age and generated a score upon completion of the questionnaire. With this EHR integration, SWYC questionnaires were seamlessly accessed and administered by providers during the visit, documented and scored directly into the chart, with an assessment as typical or delayed development. The pilot launched on November 14, 2022, with several team members testing out the questionnaire during encounters, identifying unexpected glitches, and informally tracking time required to complete and chart the questionnaires. Problems were communicated to the institution’s Epic support team and adjustments were made accordingly. An EHR tool was created in September 2023 to automatically pull SWYC questionnaire results and interpretations into the patient encounter note.

### Implementation and Interventions

Our project team officially launched the EHR-integrated SWYC questionnaire to pediatric residents and outpatient faculty members in February 2023 through targeted email blasts. All providers were instructed to administer the age-appropriate SWYC questions during the clinic encounter and complete the corresponding SWYC tab integrated into Epic. As in our previously established workflow, EI referrals were initiated when patients were identified as being at risk of developmental delay through a referral to an ambulatory social worker referral. Further interventions consisted of an additional email blast to remind providers of the implementation (March 2023), and the inclusion of SWYC screening reminders into daily team huddle announcements at the start of clinic sessions (April 2023). We had several other educational interventions over the course of our project including a residency-wide presentation and additional instructional emails in June 2023. In July 2023, a new group of pediatric residents started the program at Mount Sinai Hospital and were educated during their orientation about utilizing the developmental screening workflow (Figure 3).

**Figure 3:**
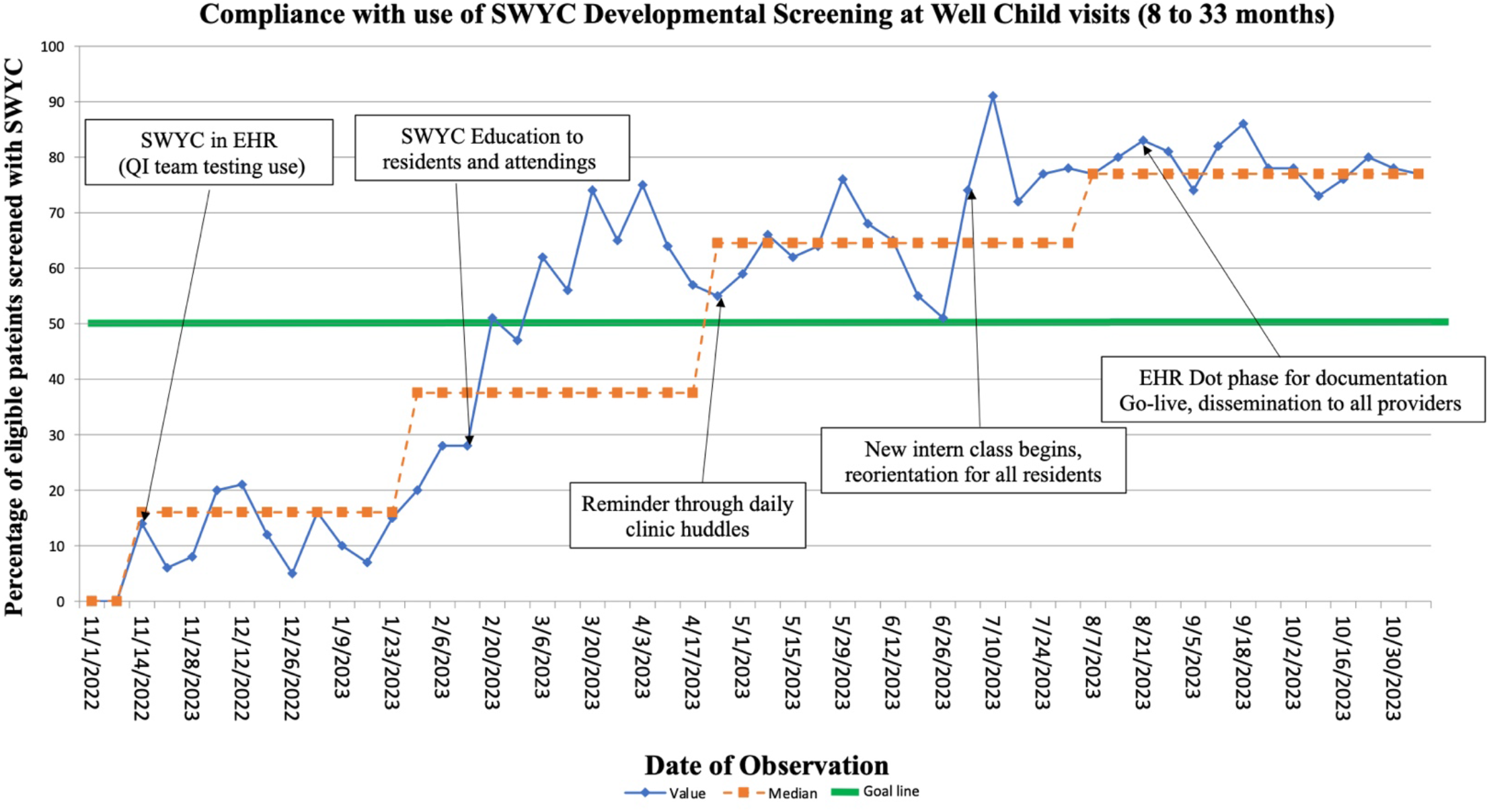
Run Chart: the SWYC Performance Trend, Annotated with Major Events.

### Data Collection and Analysis

After launching the SWYC in November 2022, our team members started manually tracking the rate of SWYC performance. Subsequently, we secured access to an automated report which included patient demographics (name, medical record number, birthdate, preferred language, and insurance coverage), as well as well-child visit date, age at visit, visit provider SWYC score and assessment and presence and date of any social worker referral. This report was sent to QI team members in an Excel spreadsheet on a monthly basis to allow for auditing of our process and outcomes and plan and implement improvement strategies.

Using the extracted Excel spreadsheet extracted by the IT team, we evaluated whether there was a significant difference in SWYC performance between patients with English as their primary language and those whose preferred language was not English. . Non-English primary language was defined based on the preferred language recorded in Epic, which is entered by patients either through the patient portal or onsite during registration. A Chi-square test was conducted using R software to analyze the data.

### Secondary Outcome Data Collection and Analysis

After implementing the SWYC and assessing the completion rates of screening questionnaires, we began to evaluate its secondary impact on patients, specifically focusing on referrals to social workers for EI. In our general pediatrics clinic, the workflow includes a designated social worker to initiate the EI referral process for any patients with concern for developmental delay.

We collected data from before and after the implementation of the SWYC to evaluate whether the rate of SW referrals (used as a proxy for EI referrals) changed significantly after the standardization of developmental screening. We extracted monthly Excel data provided by the IT team for two periods: pre-SWYC (April 2022 to November 2022) and post-SWYC (April 2023 to November 2023), when more than 50% of eligible patients underwent SWYC monthly. We excluded patients for whom a SW referral had already been made prior to the encounter, as including those patients would not accurately reflect referrals due to our interventions because each visit’s data in the Excel sheet only recorded the first date of a SW referral, which could have led to an overestimation of referral rates if the same patient visited the clinic multiple times. We calculated the number of new SW referrals divided by the total number of eligible patient encounters per month. We hypothesized that implementing a standardized screening tool would not significantly increase SW referral rates to EI services and assessed this using a Chi-square test in R.

## Results

### Result 1: SWYC Performance Rate

Before implementation, the baseline SWYC screening rate was 0%. Compliance with screening was plotted on an annotated run chart showing incremental success resulting from our interventions (Figure 3). Shifts of 8 points above initial and subsequent median lines, allowed for multiple recalculations of the median rates: from 16% (Nov 2022-Jan 2023), to 37.5% (Feb 2023-Apr 2023), to 64.5% (May 2023-Jul 2023), and finally to 77% (Aug 2023-Oct 2023). Compliance first exceeded our goal of 50% four months after the implementation of the SWYC.

### Result 2: Equitable Screening Practices for English Primary Speakers and Non-English Primary Speakers

A Chi-square test conducted using R software yielded a p-value of 0.13 (Odds Ratio 1.27, 95% confidence interval 0.93-1.72), indicating no significant difference in SWYC implementation between English primary and non-English primary speakers. (**Table 2**) Thus, we achieved the project’s goal of ensuring equitable screening practices for both English-speaking patients and those for whom English is not the primary language.

**Table 2:** Comparison of SWYC Performance Between English Primary and Non-English Primary Speakers from April to November 2023, and Their Significance.

|  | **SWYC**<br>**Performed** | **SWYC**<br>**Non-Performed** | ***P*** |
| --- | --- | --- | --- |
| **English speaker (2023 Apr-Nov)** | 1219 | 405 | 0.13 |
| **Non-English primary speaker (2023 Apr-Nov)** | 173 | 73 |  |

### Result 3: Social Worker Referral Rate

Our EHR data collection also included tracking of social worker referrals. **Figure 4** shows the total SW referral rate from April to November for both 2022 (pre-SWYC) and 2023 (post-SWYC). The pre-SWYC SW referral rate was 9.6%, and the post-SWYC referral rate increased to 12.5%. The referral rate increased by 34%. The resulting p-value of 0.009 (OR=1.3, 95% confidence interval: 1.1–1.7) indicates a statistically significant increase in referrals, supporting the alternative hypothesis of an association. (**Table 3**).

**Table 3:** Number of Social Worker (SW) Referrals Pre- and Post-SWYC from April to November 2023, and the Fisher’s Exact Test P value.

|  | **Before SWYC (2022 Apr-Nov)** | **After SWYC (2023 Apr-Nov)** | ***P*** |
| --- | --- | --- | --- |
| **SW Referral +** | 157 | 217 | 0.00854 |
| **SW Referral –** | 1484 | 1524 |  |

**Figure 4:**
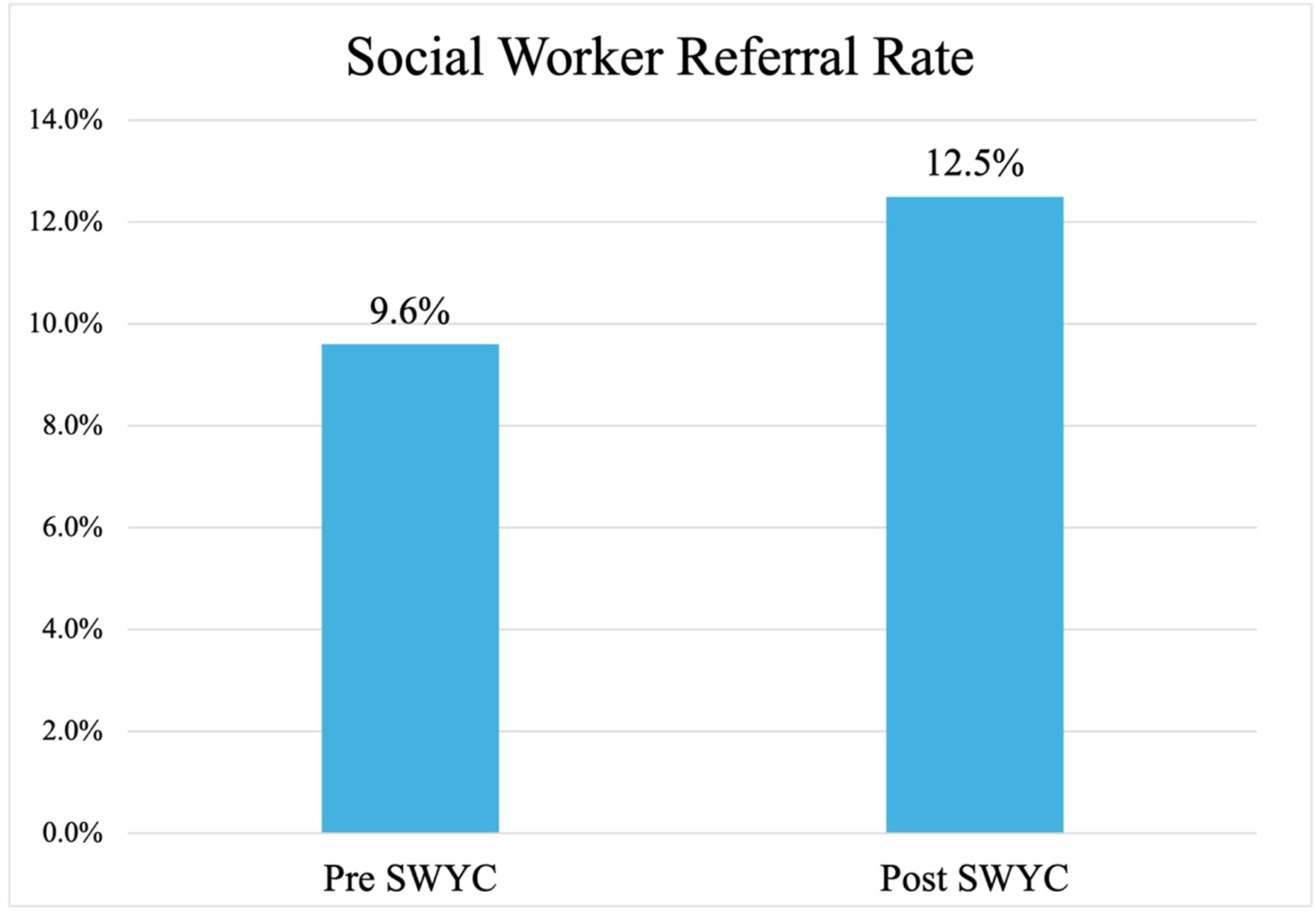
Social Worker Referral Rate of Pre- and Post-SWYC implementation

## Discussion

The AAP survey data revealed that 37% of pediatricians did not perform developmental screenings, and 41% of their at-risk patients were not referred to EI.^11^ Due to the improved therapy outcomes and better developmental progress associated with EI ^3-7^, early identification of developmental delays through screening during well-child visits and connecting children to EI services, such as occupational, physical, and speech therapy,^25^ to enhance long-term outcomes, is crucial. However, implementing a new developmental screening system within a healthcare framework is complex due to the involvement of numerous stakeholders, such as providers, nurses, medical assistants, administrative staff, IT teams, and patients and families.^26^ Challenges to success include limited time for patient encounters, the need for updating professional knowledge, and the difficulty of integrating new practices into an existing culture in clinical spaces.

Our team completed a preparatory phase involving extensive research and planning, followed by collaboration with our institution’s EHR support team to integrate the SWYC screening questionnaires into the EHR. This integration allowed for the official launch of our project, with clinical screenings beginning in November 2022. Following the initial EHR setup, through iterative PDCA cycles with numerous interventions we expanded the implemented new screening process to all general pediatric providers in our clinic site. Interventions included emails to all ambulatory providers, oral presentations, daily huddles, and verbal reminders, leading to a swift adaptation and a rapid increase in its use. We achieved our goal of over 50% within four months of launch with equitable screening practices for both English-speaking patients and those for whom English is not the primary language. Following the implementation of SWYC, the referral rate to social workers increased by 34%.

The success of our QI project can likely be attributed to the system’s ease of use, seamless integration into the EHR (eliminating the need for paper forms or manual entry), a practical scoring system that provides both scores and management recommendations, and early buy-in from stakeholders. We surmise that the increase in EI referrals is due to the practical scoring and management recommendations integrated with the SWYC in our EHR. These features support clinicians’ judgment based on history taking and patient observation, potentially increasing their confidence in referring patients to EI.

There are several limitations in this study. We cannot state as a fact that all the social work referrals were exclusively for EI referrals since other needs also arise - for example, housing concerns, and food insecurity. However, based on a review of the practice’s social work referrals, we know that most referrals for children aged 0–3 years were for EI evaluations, and our clinic’s workflow, which includes a designated social worker to initiate the EI referral process, makes this outcome a reliable proxy. For sustaining the project, it is crucial to continue to reiterate the SWYC screening to clinicians and monitor screening performance rates to improve compliance rates towards 100%.

## Conclusions

A new validated developmental screening tool utilizing the SWYC was successfully implemented in a busy residency continuity clinic and was seamlessly integrated into the EHR, allowing for objective scoring and interpretation. The utilization of this tool has likely enabled the timely detection of developmental delays and enhanced the facilitation of appropriate referrals to social workers for EI evaluations. Implementing a standardized developmental screening system holds significant potential for improving developmental outcomes for children in the U.S. and enhancing care equity through an objective scoring and assessment system.

## Data Availability

All data produced in the present study are available upon reasonable request to the authors.

